# Outcome of Different Therapeutic Interventions in Mild COVID-19 Patients in a Single OPD Clinic of West Bengal: A Retrospective study

**DOI:** 10.1101/2021.03.08.21252883

**Authors:** Sayak Roy, Shambo Samrat Samajdar, Santanu K Tripathi, Shatavisa Mukherjee, Kingshuk Bhattacharjee

## Abstract

**Introduction:** With over 87,273,380 cases being reported and 1,899,440 deaths worldwide as of 9th January 2021, Coronavirus disease 2019 (COVID-19) has become the worst-hit pandemic till date. Every day clinicians are bombarded with many new treatment options that claim to be better than the others.

**Materials and methods:** After screening the electronic database of COVID-19 patients retrospectively, 56 patients with mild COVID-19 infection matched the inclusion criteria and were divided into the four following groups - group having used Hydroxychloroquine (HCQ), group using doxycycline (DOX) + Ivermectin (IVR) combination, group receiving only azithromycin (AZ) and, group receiving only symptomatic treatment. The study’s primary objective was to see Clinical response of well-being (CRWB) reporting time after initiating treatment onset between the four different treatment arms.

**Results:** CRWB did not differ between the four groups receiving four different managements (p-value 0.846). There was significant correlation between blood levels of LDH (p-value 0.001), CRP (p-value 0.03) and D-dimer (p-value 0.04) with CRWB in IVR+DOX group and, between LDH (p-value 0.001), CRP (p-value 0.01) and age (p-value 0.035) with CRWB in the symptomatic management group.

**Conclusion:** Mild COVID-19 infection in patients having low-risk to progress can be managed symptomatically without any specific drug intervention.

## Introduction

As of 9th January 2021, there has been a worldwide record hit of 87,273,380 cases and 1,899,440 deaths due to the COVID-19 [**1**]. Guidelines have been quite variable when it comes to treating mild COVID-19 infections, from only symptomatic treatments as per National Institutes of Health (NIH) [**2**] to IVR+DOX or HCQ in others [**3**]. These varied recommendations are always creating confusion amongst physicians. There is no universally accepted or recommended guideline on mild COVID-19 infection treatment.

This study will throw some light by comparing these four groups with all the recommended medications as per guidelines and their outcome.

## Materials and methods

The present study considered all SARS-CoV-2 RT-PCR positive patients having mild symptom onset of less than three days, visiting the concerned OPD (out-patient department) clinic from 5th April 2020 to 11th January 2021. Their medical data were retrieved from the electronic database (EDB) of the investigator after taking written consent. Patients requiring oxygenation or, admission due to any cause were excluded from the study. This study was approved by an Institutional ethical committee and followed the declarations laid down in Helsinki’s declaration. The patients were divided into four groups – 1) having HCQ 200 mg twice daily (400 mg on Day 1 only), 2) IVR+DOX (IVR as once-daily of 12 mg and DOX 100 twice daily), 3) AZR 500 mg once daily, and, the last group as per NIH guidelines who refused any treatment other than supportive treatment with antipyretics and oral rehydration solution.

The study’s primary objective was to see Clinical response of well-being (CRWB) reporting time after initiating treatment onset between the four different treatment arms. Blood tests for D-dimer, Neutrophil to lymphocyte ratio (NLR), C-reactive protein (CRP) and, Lactate dehydrogenase (LDH) were done after their first visit at the clinic within the second day after starting treatments. Blood for IgG SARS-COV-2 was done after 21 days to check the antibody titer. All the patients were under telephonic supervision for 14 days as per home isolation guidelines after their first initial turn up in the clinic [**4**]. Informed consent forms were distributed and collected from participants who fell under the case definition of mild COVID-19 infection, after initial screening of the EDB of *mild COVID-19 patient category* from the investigator’s data base. Enrolled patients were interviewed regarding any existing comorbidities. All of these patients received vitamin C & D and oral zinc supplements throughout these 14 days.

## Statistical analysis

Since there is lack of evidence on this type of studies, we took a minimum of 30 samples as a rule of thumb. All data were captured by the treating physician during treatment and recorded electronically. Data were retrieved retrospectively from the EDB of these patients. Data collected were checked for completeness and analyzed using statistical software, namely Statistical Package for Social Sciences (SPSS Complex Samples) Version 21.0 for Windows, SPSS, Inc., Chicago, IL, USA, along with MS Excel. We performed a one-way Analysis of variance (ANOVA) to see any statistical significance between CRWB and various baseline parameters in the different treatment arms. We also performed Spearman’s Correlation test (2-tailed) to see the correlation between various parameters with CRWB in the different arms.

## Result

A total of 73 patients with mild COVID-19 were initially screened, out of which 56 met the baseline inclusion criteria and consented to be a part of the study. The baseline demographics are given in Table 1.

**Table 1.**
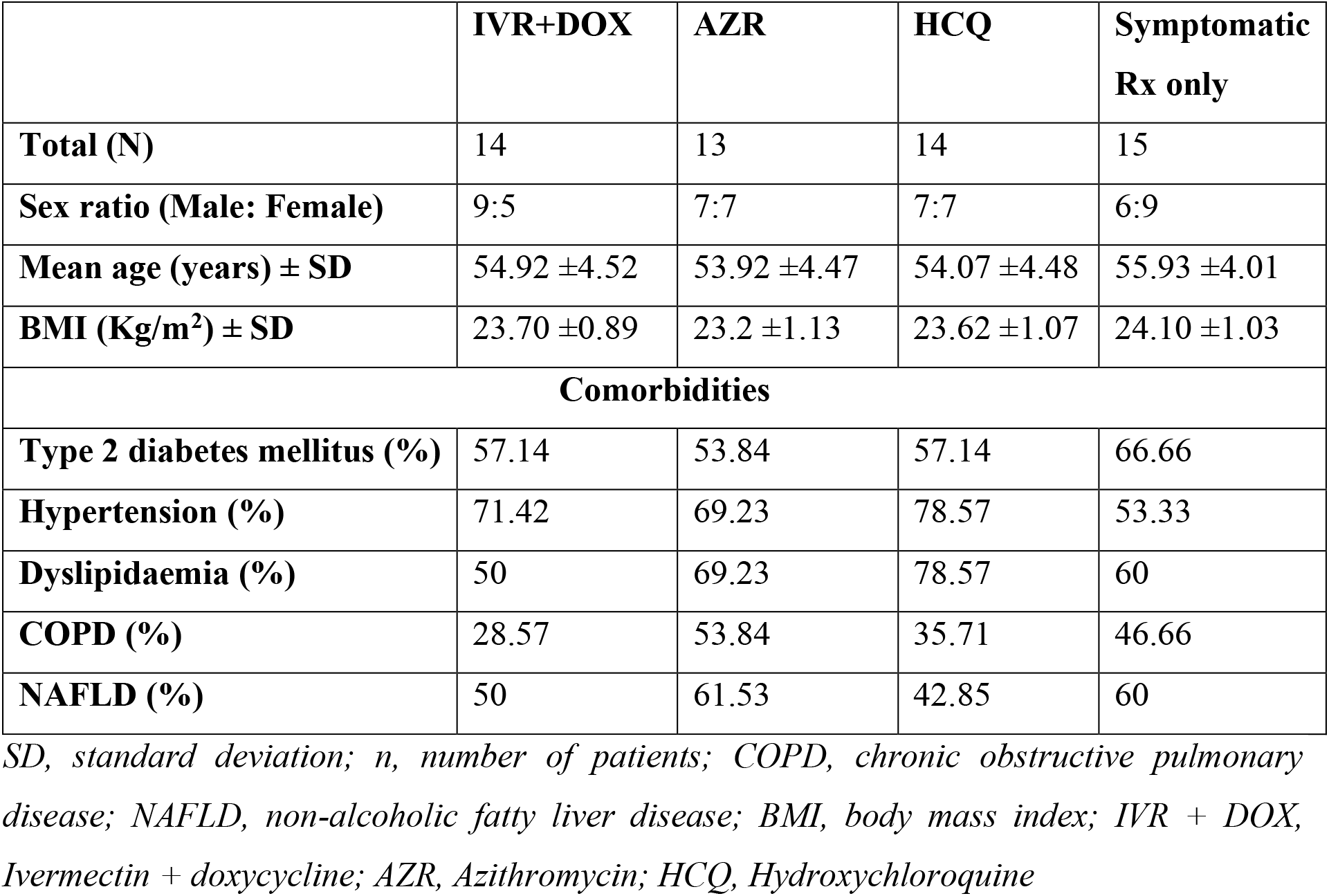
Baseline demographics.

The results of all the various treatments showed no difference, as measured by ANOVA, in outcome in terms of “Clinical response of wellbeing (CRWB),” as described by the patient of a feeling of wellbeing with subsidence of fever and, spearman’s correlation test was also done to see any correlation between various parameters with CRWB of each group. The ANOVA also showed no difference in the various baseline parameters that could have impacted our various treatment protocols and outcomes. They are all summarised in Table 2.

**Table 2.** Analysis of various parameters with CRWB.

| **ANOVA analysis of various parameters** |  |  |  |  |  |
| --- | --- | --- | --- | --- | --- |
| **Parameters (Mean, SD)** | **IVR + DOX**<br>**N = 14** | **AZR**<br>**N = 13** | **HCQ**<br>**N = 14** | **Symptomatic Rx**<br>**N = 15** | **P value** |
| **Age** | 54.93 ± 8.96 | 53.92 ± 8.57 | 54.07 ± 8.87 | 55.93 ± 8.20 | 0.92 |
| **BMI** | 23.71 ± 1.77 | 23.2 ± 2.16 | 23.62 ± 2.13 | 24.10 ± 2.1 | 0.71 |
| **LDH** | 230.43 ± 41.49 | 215.61 ± 33.58 | 226.71 ± 53.86 | 232.73 ± 52.22 | 0.78 |
| **CRP** | 1.15 ± 0.94 | 2.31 ± 3.64 | 1.96 ± 2.36 | 1.33 ± 0.89 | 0.485 |
| **D-dimer** | 0.45 ± 0.04 | 0.50 ± 0.12 | 0.49 ± 0.08 | 0.50 ± 0.10 | 0.526 |
| **NLR** | 3.24 ± 1.0 | 3.42 ± 1.55 | 3.28 ± 0.76 | 3.36 ± 1.12 | 0.976 |
| **CRWB (days)** | 3.21 ± 0.38 | 3.23 ± 0.78 | 3.32 ± 0.61 | 3.4 ± 0.69 | 0.846 |
| **IgG value\*\*** | 6.49 ± 2.17 | 6.09 ± 2.61 | 6.23 ± 2.47 | 6.70 ± 2.66 | 0.92 |
| **Spearman's Correlation test between each group with CRWB** |  |  |  |  |  |
| **Parameters** | **P-value**<br>**(2-tailed)** | **IVR + DOX** | **AZR** | **HCQ** | **Symptomatic**<br>**Rx** |
| *NLR*: |  | 0.303 | 0.29 | 0.577 | 0.437 |
| *LDH* |  | 0.001* | 0.07 | 0.601 | 0.001* |
| *CRP* |  | 0.03* | 0.22 | 0.203 | 0.01* |
| *D-dimer* |  | 0.04* | 0.103 | 0.521 | 0.284 |
| *AGE* |  | 0.07 | 0.557 | 0.919 | 0.035* |
| *BMI* |  | 0.311 | 0.479 | 0.08 | 0.456 |

\*statistically significant at $P$ -value $<0.05$ ; \*\*Negative taken as Index $<1.0$ ; SD, standard deviation; $n$ , number of patients; COPD, chronic obstructive pulmonary disease; BMI, body mass index; IVR + DOX, Ivermectin + doxycycline; AZR, Azithromycin; HCQ, Hydroxychloroquine; NLR, Neutrophil to lymphocyte ratio; LDH, Lactate dehydrogenase; CRWB, Clinical response of wellbeing

## Discussion

Mild COVID-19 is defined by confirmed COVID-19 case having no evidence of viral pneumonia or hypoxia [**5**]. Mild COVID-19 treatment has been bombarded with almost all sorts of medicines from all classes, starting from high dose famotidine [**6**] to anti-parasitic agent Ivermectin [**7**] to inhaled nitrous oxide [**8**]. The guidelines are formed depending on the various ongoing trials and their results. Ivermectin is an anti-parasitic drug that has been used for mild SARS-COV-2 treatment since it is presumed to block viral proteins from entering the host cell nucleus [**7**]. Doxycycline is thought to block SARS-CoV-2 papain-like protease enzyme [**9**]. Azithromycin with hydroxychloroquine has been used in many studies for COVID-19 treatment with contradictory results [**10**]. Azithromycin seems to have multiple modes of actions through which it affects the SARS-COV-2 virus-like, anti-inflammatory property, antiviral effects in bronchial epithelial cells and, immunomodulatory property [**11**]. Hydroxychloroquine has been found to have an antiviral effect when used in-vitro [**12**]. All these innumerable small scales and large randomized trials have led to the formation of numerous guidelines recommending various drugs that can be used in mild COVID-19 infection. Guidelines are highly variable, with Singapore asking to use supportive therapy only in low-risk group having non-severe COVID-19 and using lopinavir/ritonavir or subcutaneous Interferon beta-1B or HCQ or Remdesivir on a trial basis in high-risk non-severe cases [**13**]; world health organization (WHO) recommending only symptomatic treatments for mild infection [**5**]; CDC recommending SARS-CoV-2 neutralizing antibodies available through emergency use authorization (bamlanivimab or, acasirivimab plus imdevimab) only for patients having risk for progression [**14**] and, India recommending supportive therapy to use of IVR or, HCQ as per patient [**15; 3**]. Studies have shown benefit with IVR (NCT04422561) as post-exposure prophylaxis (within 72 hours), but when we study in-depth, we find that the secondary outcome of confirmation of positivity by RT-PCR was not reported, and the clinical history was only taken as a benchmark of getting COVID-19 infection [**16**]. Another trial of IVR (NCT04425850), done on health personnel, have shown good protective results with buccal drops of IVR given to health personnel on top of standard of safety to be followed regularly, and it came up with a brilliant 0% RT-PCR positive cases in IVR arm as compared to 11.2% positive in standard arm [**17**]. This study has the drawbacks of being an observational, prospective trial with no blinding again. Many guidelines are enumerating many drugs for mild COVID-19 patients, where mild infection is defined by the presence of mild upper respiratory symptoms in an uncomplicated severe acute respiratory syndrome coronavirus 2 (SARS-CoV-2) reverse transcription-polymerase chain reaction (RT-PCR) positive patient [**5**]. To assess these varied treatment protocols in mild COVID-19, this retrospective case study was done to see any change in clinical wellbeing reporting onset timing difference in patients receiving three of the presently available protocols and comparing that to the group who declined any treatments except symptomatic management only. A study on 74 hospitalized patients showed an NLR of >4 to be associated with admission to ICU and young age, as well as an NLR of <3 was associated with clinical improvement [**18**]. Low levels of lymphocytes, high serum LDH, ferritin, D-dimer are often associated with poor outcomes in COVID-19 [**19**]. Blood CRP level amongst hospitalized patients of COVID-19 showed a correlation with the disease outcome, with survivors having a median value of 40 mg/L and non-survivors having a median CRP level of 125 mg/L [**20**].

This study showed no change in clinical wellbeing reporting onset timing between all the groups (p-value 0.846). There was also no significant difference between the baseline parameters, as measured by ANOVA, which could have affected the outcome. This study showed positive significant correlation between blood levels of LDH (p-value 0.001), CRP (p-value 0.03) and D-dimer (p-value 0.04) with CRWB in IVR+DOX group and, between LDH (p-value 0.001), CRP (p-value 0.01) and age (p-value 0.035) with CRWB in the symptomatic management group. The other two groups did not show any correlation between CRWB and any other parameters. The small sample size, coupled with the study’s retrospective nature, could not allow us to investigate further into the reason for this finding. However, our study is not without limitations. A record-based study has less power to draw any conclusion and comes with few biases. Moreover, the small sample size and single-center set-up hinders data generalizability to an extent. It was even difficult to state whether patients’ followed instructions or not and was based on their telephonic response, which invites bias. We could not perform serial RT-PCR to confer on patients’ decreasing viral load.

## Conclusion

Tons of publications on COVID-19 have posed a severe problem in choosing the correct treatment for mild COVID-19 infection. Guidelines are trying to find new therapeutic strategies to tackle this new infection, which is only one year old. Many new strains of COVID-19 are emerging with varying modes of presentations, increasing physicians’ dilemma to a greater extent. Mild COVID-19 infections can be managed symptomatically in low-risk groups. Only the high-risk groups can be taken into consideration of interventions. This study can help us understand that we might use symptomatic management in mild COVID-19 infections in those who do not progress. These findings need to be further investigated in a larger sample size to reach any conclusion. Case series like this can only give us an idea and, a conclusion regarding treatment protocols in mild infections must be made after doing large-scale prospective, randomized studies. To ease physicians’ dilemma, there must be one universally accepted treatment protocol for these mild infections.

## Data Availability

Available with the corresponding author on request.

## Acknowledgement

None

## Notes

**Conflicts of interest:** None

**Funding:** None

### Competing Interest Statement

The authors have declared no competing interest.

### Author Declarations

This study was reviewed by the Institutional ethical committee of the School of Tropical Medicine, Kolkata (IEC Ref No: CREC-STM/2020-AS-36) and clearance was granted. The methods were followed as per the Declaration of Helsinki.

### Summary of Updates

The consent part of "Materials and methods" section of page 3 has been modified.

